# Epigenetic Clock Trajectories and Brain Health in Midlife

**DOI:** 10.64898/2026.07.16.26358251

**Authors:** Ana I. Boeriu, Shea J. Andrews, Tina Hoang, Sejong Bae, Kristine Yaffe

## Abstract

**Background:** Accelerated biological aging can be assessed with DNA methylation (DNAm)-based epigenetic clocks. Research suggests that greater DNAm is associated with faster cognitive decline and risk of Alzheimer disease (AD) and other dementias. However, most studies have relied on single-time-point measurements of clocks, rather than evaluating dynamic changes over time. We examined the association between 15-year epigenetic aging trajectories and brain health outcomes in midlife.

**Methods:** We analyzed 2,833 middle-aged adults (mean baseline age 40 years, 59% female and 44% Black) with ≥ 3 DunedinPACE (a recently developed epigenetic clock) measurements, collected over 15 years. Using mixed-effects modeling, we derived individual-specific slopes of epigenetic aging trajectories and categorized participants as Fast Agers (slopes > 1 SD above the mean), Slow Agers (slopes < 1 SD below the mean), or Typical Agers (within ±1 SD of the mean). We examined associations between trajectory group and cognition on five cognitive domains as well as on plasma AD biomarkers (NfL, p-tau217, Aβ42/Aβ40), all assessed 15-20 years post-baseline. Models were adjusted for demographics, education, physical activity and *APOE*\*ε4 carrier status (with additional adjustments for eGFRcr for biomarker outcomes).

**Results:** Epigenetic aging trajectories were associated with multiple domains of cognition and AD biomarkers (Figure 1). Compared to Typical Agers, Fast Agers showed worse processing speed, memory, executive function, and global cognition (all p<0.05), with no difference in verbal fluency. Slow Agers had better performance on memory and global cognition (both p < 0.05). Fast Agers also exhibited significantly lower Aβ42/Aβ40 levels (p = 0.011) compared to Typical agers; no significant associations with p-tau217 or NfL were observed in either group.

**Conclusion:** Middle-aged adults with faster 15-year epigenetic aging trajectories demonstrated worse cognitive performance, whereas those with slower biological aging trajectories exhibited cognitive resilience and more favorable AD biomarker profiles. By examining long-term trajectories rather than single timepoints, these findings identify individuals at differential risk for brain health outcomes.

## Introduction

Not all individuals age at the same rate. While chronological age advances uniformly, biological aging—the progressive decline in physiological function—varies substantially across individuals due to genetics, lifestyle, and environmental factors.

**Figure 1:**
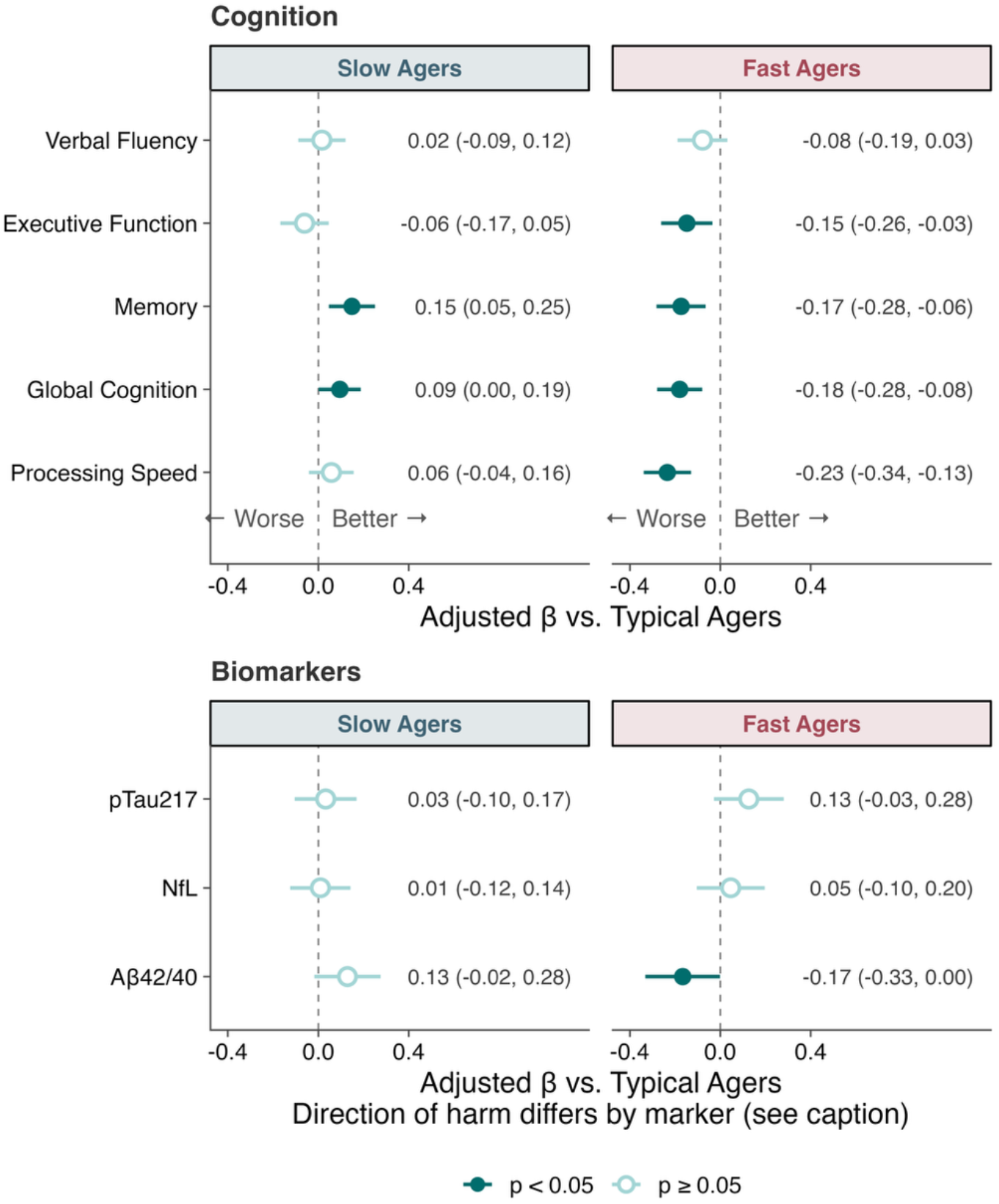
Adjusted associations of epigenetic aging trajectory groups with cognitive performance and plasma AD biomarkers. Models were adjusted for age, sex, race, education, physical activity, *APOE*\*ε4 carrier status, and, additionally, for eGFR (biomarker outcomes). Point estimates represent unstandardized regression coefficients (β) with 95% confidence intervals comparing Slow Agers and Fast Agers to Typical Agers (reference group). Cognitive outcomes are z-scored; biomarkers are log10-transformed and z-scored. Dark points indicate p < 0.05

Epigenetic clocks leverage DNA methylation patterns to quantify biological aging independent of chronological aging^1,2^, capturing molecular changes that predict mortality, morbidity, and cognitive outcomes, including the risk of Alzheimer disease^3–6^. Cross-sectional studies have linked higher epigenetic age to worse cognitive performance^7,8^, reduced brain volume^9^, and less favorable AD biomarker profiles in older adults^10^. Unlike traditional epigenetic clocks that estimate biological age, DunedinPACE quantifies the pace of aging—how fast individuals age per calendar year^5^. Developed using machine learning and trained on longitudinal biomarkers of physiological decline across multiple organ systems, a faster DunedinPACE has been associated with worse cognitive performance^11^, lower hippocampal volume, and a thinner cerebral cortex^12^. However, DunedinPACE studies have relied on single-time-point measurements, which cannot distinguish individuals with sustained trajectories of accelerated aging from those with temporary fluctuations.

Longitudinal trajectories of epigenetic aging may provide stronger predictions of brain health outcomes by capturing the rate at which biological processes unfold, rather than one-time snapshots^13,14^. In addition, most prior research has focused on older adults, potentially missing critical periods when aging trajectories begin to diverge. The midlife period is a particularly important developmental window during which early pathological changes linked to AD may emerge without noticeable cognitive impairment^15^. The association between epigenetic aging trajectories during midlife and brain health outcomes remains largely unexplored. By focusing on midlife when cognitive function is typically stable, yet dementia pathology may be emerging, we can identify individuals at elevated risk while interventions may still modify disease trajectories.

We derived 15-year (baseline age, mean = 40.31, range = [32,49]) epigenetic aging trajectories using DunedinPACE from DNA methylation measurements collected across four time points in middle-aged adults enrolled in the Coronary Artery Risk Development in Young Adults Study (CARDIA) and characterized participants as slow, typical, or fast agers based on their rates of biological aging over time. We next examined associations between aging trajectory groups and cognitive performance across five domains, as well as plasma AD biomarkers (Aβ42/Aβ40 ratio, p-tau217, NfL) assessed 15-20 years later. We hypothesized that faster epigenetic aging trajectories would predict worse cognitive performance and less favorable biomarker profiles, while slower trajectories would indicate cognitive resilience and healthier biomarker patterns.

## Methods

### Study Population

We studied participants enrolled in the Coronary Artery Risk Development in Young Adults (CARDIA) study, a US multicenter prospective cohort study investigating cardiovascular health and aging across the life course^16^. Initially, 5,115 Black and White healthy adults aged 18-30 years were recruited from four US cities (Birmingham, AL; Chicago, IL; Minneapolis, MN; and Oakland, CA)^14,17–19^. Participants completed ten follow-up examinations, approximately every 2-5 years, with retention rates among surviving participants exceeding 70% at each examination^16^. The analytic cohort for this study included 2,833 participants with DunedinPACE measurements for at least 3 out of 4 possible time points across a 15-year period, spanning from the CARDIA year 15 clinic visit (our study baseline) to year 30 visit. Cognitive outcomes were assessed 15 or 20 years after baseline, and plasma biomarkers were assessed 20 years after. All participants provided written informed consent at each examination center, and study protocols were reviewed by institutional review boards at each study site and the CARDIA Coordinating Center^18^.

### DNA Methylation/DunedinPACE

DNA methylation was measured using Illumina’s Infinium MethylationEPIC Bead Chip (~850,000 CpGs) in the TOPMed program^14^. DunedinPACE scores were computed using an established algorithm and implemented in R using publicly available code^5^.

DunedinPACE quantifies the rate of biological aging rather than biological age at a single time point^5^. The criterion phenotype, Pace of Aging, was derived from longitudinal modeling of within-person change across 19 biomarkers of cardiovascular, metabolic, renal, immune, dental, and pulmonary organ-system integrity assessed at ages 26, 32, 38, and 45^5^. DunedinPACE was then trained via elastic-net regression to predict the 20-year Pace of Aging. Candidate probes were restricted to CpG sites shared by the Illumina 450K and EPIC arrays with acceptable test–retest reliability, yielding a 173-CpG algorithm^5^. Values are scaled such that 1.0 reflects one year of physiological decline per chronological year, with higher values indicating faster aging^5^.

### Epigenetic Aging Trajectory Categorization

To characterize individual trajectories of epigenetic aging over the 15-year period, we fit a linear mixed-effects model with random intercepts and random slopes, where time was coded as years since baseline. From this model, we extracted individual-specific slopes representing each participant’s rate of DunedinPACE change over the 15-year period (Supplementary Figure 1). Individual slopes were standardized (z-scores), and participants were categorized as Fast Agers (slopes > 1 SD above the mean), Slow Agers (slopes < 1 SD below the mean), or Typical Agers (within ±1 SD of the mean) (Supplementary Figure 2).

### Cognitive Assessment

Cognitive performance was assessed at 15 or 20 years post baseline using a neuropsychological battery consisting of the Montreal Cognitive Assessment (MoCA; global cognition), Rey Auditory Verbal Learning Test delayed recall (RAVLT; verbal memory), Digit Symbol Substitution Test (DSST; processing speed), Stroop Interference Test (executive function), and Letter and Category Fluency (verbal fluency)^20^. All cognitive test scores, including Stroop, were coded so that higher values indicate better performance. Total verbal fluency was calculated as the sum of letter and category fluency scores.

### AD Biomarker Measurements

AD biomarker assays were performed among participants who attended the 20-year post-baseline examination and had 15–20 years post-baseline cognitive assessments. Blood samples were processed and stored at −70 °C within 90 minutes until assayed at the Laboratory of Pathology and Laboratory Medicine at the Hospital of the University of Pennsylvania. Plasma p-tau217 and Aβ^42^/Aβ^40^ were assayed using Fujirebio Lumipulse G1200 analyzer as previously described^15^.

### Covariates

Covariates were selected based on known associations with epigenetic aging, cognitive function, and brain health, as well as previous literature in CARDIA. Baseline covariates included age, sex, race, education (years), physical activity and *APOE\**ε4 carrier status. Physical activity was measured using the validated CARDIA Physical Activity History questionnaire^21,22^. Domain-specific covariates for plasma biomarker included estimated glomerular filtration rate (eGFRcr) calculated using the 2021 CKD-EPI creatinine equation without race^23^. *APOE* genotype was dichotomized as ε4 carrier (≥1 ε4 allele) versus non-carrier, with ε2/ε4 heterozygotes classified as ε4 carriers given the established risk associated with the ε4 allele^24^.

### Statistical Analysis

Descriptive statistics compared participant characteristics by epigenetic aging trajectory group using χ^2^ tests (categorical variables) and ANOVA (continuous variables). We used linear regression to examine associations between aging trajectory groups (Fast Agers, Slow Agers vs Typical Agers) and cognitive performance and biomarker outcomes. Linear regression models adjusted for demographics, education, and physical activity, and *APOE\**ε4 carrier status. Model fit was summarized using R^2^/ICC for the mixed model and adjusted R^2^ for outcome models. As a sensitivity analysis, we additionally adjusted for baseline epigenetic aging defined as the model-estimated DunedinPACE at baseline (sum of the fixed and random intercepts). We also evaluated two-way interactions between epigenetic aging and sex, race, and *APOE*\*ε4 carrier status in adjusted models. Analyses were conducted using R version 4.5.2. Statistical significance was defined at α= 0.05.

## Results

### Epigenetic Aging Trajectories

Between-person differences accounted for most of the variance in DunedinPACE (ICC = 0.77; conditional R^2^ = 0.77) and including a random slope for time significantly improved fit over a random-intercept-only model (χ^2^(1) = 283.8, p < 0.001), consistent with meaningful individual variation in the rate of change.

### Study Population

Among 2,833 participants with epigenetic trajectory data, the mean age at baseline was 40.3 ± 3.6 years, 58.5% were female, and 44.0% were Black (Table 1). Based on individual epigenetic aging slopes over 15 years, 404 (14.3%) participants were classified as Slow Agers, 2,023 (71.4%) as Typical Agers, and 406 (14.3%) as Fast Agers. Slow Agers were more likely to be female, White, and have more education (p < 0.05 for all) (Table 1).

**Table 1:** Baseline Characteristics by Epigenetic Aging Trajectory.

| **Characteristic** | **Overall** | **Slow Agers** | **Normal Agers** | **Fast Agers** | **p-value<sup>2</sup>** |
| --- | --- | --- | --- | --- | --- |
|  | N = 2,833<sup>1</sup> | N = 404<sup>1</sup> | N = 2,023<sup>1</sup> | N = 406<sup>1</sup> |  |
| **Age** | 40.3 (3.6) | 40.8 (3.4) | 40.3 (3.6) | 39.8 (3.8) | 0.004 |
| **Female** | 1,658 (58.5%) | 304 (75.2%) | 1,169 (57.8%) | 185 (45.6%) | <0.001 |
| **Race** |  |  |  |  | <0.001 |
| White | 1,587 (56.0%) | 286 (70.8%) | 1,135 (56.1%) | 166 (40.9%) |  |
| Black | 1,246 (44.0%) | 118 (29.2%) | 888 (43.9%) | 240 (59.1%) |  |
| **Education (years)** | 15.1 (2.5) | 15.7 (2.5) | 15.0 (2.5) | 14.6 (2.5) | <0.001 |

<sup>1</sup>Includes all participants with epigenetic trajectory data (N=2,833).
<sup>2</sup>One-way ANOVA; Pearson's Chi-squared test

### Epigenetic Aging Trajectories and Cognitive Performance

In the unadjusted analysis with Typical Agers as reference, Fast Agers showed significantly worse cognitive performance across all cognitive domains, including processing speed (β = −0.39; 95% CI: −0.5, −0.27), memory (β = −0.31, 95% CI: −0.42, −0.20), global cognition (β = −0.36, 95% CI: −0.48, −0.25), executive function (β = −0.26, 95% CI: −0.38, −0.15), and verbal fluency (β = −0.22, 95% CI: −0.33, −0.11; all p < 0.05). Slow Agers showed significantly better performance on processing speed (β = 0.30, 95% CI: 0.20, 0.41), memory (β = 0.37, 95% CI: 0.26, 0.48), global cognition (β = 0.32, 95% CI: 0.21, 0.42), and verbal fluency (β = 0.19, 95% CI: 0.08, 0.30; all p < 0.05), but not executive function (β = 0.11, 95% CI: −0.002, 0.21, p = 0.058)

After adjusting for age, sex, race, education, physical activity, and *APOE*\*ε4 carrier status, associations remained significant across most domains (Figure 1). Compared to Typical Agers, Fast Agers demonstrated significantly lower performance in processing speed, memory, executive function, and global cognition (βs ranging from −0.23 to −0.15, 95% CIs spanning −0.34 to −0.03; all p < 0.05). No significant association was observed for verbal fluency. Slow Agers had better performance on memory and global cognition (βs = 0.15 and 0.09, 95% CIs ranging from 0.001 to 0.25; both p < 0.05), but not on processing speed, executive function, or verbal fluency.

### Epigenetic Aging Trajectories and AD Plasma Biomarkers

Epigenetic aging trajectories were associated with plasma AD biomarkers. In unadjusted analyses, Fast Agers showed significantly lower Aβ42/Aβ40 ratios (β = −0.19, 95% CI: −0.35, −0.03) and Slow Agers showed higher Aβ42/Aβ40 ratios (β = 0.16, 95% CI: 0.01, 0.30) compared to Typical Agers. No significant associations were observed for NfL or p-tau217. After adjusting for age, sex, race, education, physical activity, eGFR, and *APOE\**ε4 carrier status, Fast Agers continued to show significantly lower Aβ42/Aβ40 ratios (β = −0.22, 95% CI: −0.40, −0.05; p = 0.011). No significant associations were observed for NfL or p-tau217 after adjustment. The Slow Agers association with Aβ42/Aβ40 was attenuated and no longer significant after adjustment. After adjustment for baseline epigenetic aging, all associations retained their direction and significance except the Slow-Ager association with global cognition, which was borderline in the primary model (β = 0.09, p = 0.047) and no longer significant after adjustment (β = 0.08, p = 0.097).

Adjusted R^2^ for the fully adjusted models ranged from 0.08 to 0.14 for plasma biomarkers and 0.15 to 0.33 for cognitive outcomes (Supplementary Table 1), with all models significant overall (all p < 0.001). No consistent interactions between aging trajectory, sex, race, or *APOE*\*ε4 status were observed across cognitive or biomarker outcomes.

## Discussion

In this study of almost three thousand middle-aged adults from the CARDIA cohort, we found that 15-year epigenetic aging trajectories derived from DunedinPACE were associated with cognitive performance and plasma AD biomarkers assessed 15–20 years later. After multivariate adjustment, Fast Agers showed significantly worse performance in processing speed, memory, global cognition, and executive function, as well as significantly lower Aβ42/Aβ40 ratios compared to Typical Agers. No significant associations were observed for verbal fluency, NfL, or p-tau217. Slow Agers demonstrated better memory and global cognition, and a trend toward higher Aβ42/Aβ40 levels. These findings suggest that sustained patterns of accelerated epigenetic aging in midlife are independently associated with worse brain health outcomes.

Across multiple epigenetic clocks, accelerated biological aging has been consistently associated with worse primarily late-life cognitive performance^7,8,25^. Within CARDIA, greater midlife epigenetic age acceleration, measured by GrimAge at year 20, was associated with worse processing speed and verbal memory five years later^26^. Using DunedinPACE clocks, a greater aging pace was associated with worse baseline cognition and more rapid decline in the Framingham Heart Study^11^, and with reduced hippocampal volume and cortical thickness across three cohorts of older adults^12^. These studies, however, relied on single-timepoint epigenetic measurements in predominantly older cohorts. By deriving individual aging slopes from four DunedinPACE assessments over 15 years in a midlife cohort, our findings demonstrate that sustained patterns of accelerated aging are associated with worse cognitive outcomes well before the typical age of onset of cognitive impairment. This is consistent with multi-cohort evidence that longitudinal changes in biological aging markers, including in CARDIA, outperform single-timepoint measures in predicting health trajectories^14^.

The AD biomarker findings complement the cognitive results by implicating amyloid pathology as a potential early marker of differential biological aging. In the Women’s Health Initiative Memory Study, a higher one-time measure of DunedinPACE was associated with faster longitudinal increases in plasma p-tau181, p-tau217, NfL, and GFAP over 15 years, whereas cross-sectional associations with the Aβ42/Aβ40 ratio were observed for AgeAccelHorvath and AgeAccelPheno clocks^10^. Our finding that Aβ42/Aβ40 was the only biomarker significantly associated with aging trajectories after adjustment is consistent with evidence that amyloid dysregulation precedes neurodegeneration and tau pathology by years to decades^27^. In a midlife cohort, amyloid-related measures may represent the earliest detectable pathological signal, whereas NfL and p-tau217 may become informative only later — as demonstrated in a late life cohort where DunedinPACE was associated with longitudinal increases in both markers over 15 years^10^.

Several limitations should be considered in interpreting our study results. As a longitudinal cohort study, attrition over the 20+ year follow-up period may introduce selection bias. Participants who remained in the study may be healthier than those lost to follow-up, potentially underestimating the true associations. The linear mixed-effects model assumes a constant rate of change in DunedinPACE over 15 years, which may obscure nonlinear aging patterns. Individual-specific slope estimates may also be less stable for participants with sparse follow-up, despite partial pooling. Categorizing participants using ±1 SD thresholds is statistically convenient but biologically arbitrary. This may misclassify individuals near cut points and result s in some loss of information compared with the continuous slope. The composition of the CARDIA cohort may also limit generalizability to broader populations. This study also has several strengths. Repeated DunedinPACE measurements across four time points over 15 years enabled individual-level trajectory characterization. Single measurements cannot distinguish persistently accelerated aging from short-term fluctuations; by deriving individual slopes from repeated assessments, our novel approach captures sustained patterns of biological aging that may more reliably predict downstream outcomes. We were also able to study a more diverse midlife population than most epigenetic aging studies, which have been conducted predominantly in older White cohorts^11,28^. The examination of both multi-domain cognitive performance and plasma AD biomarkers provides converging evidence linking epigenetic aging trajectories to brain health.

In conclusion, 15-year epigenetic aging trajectories in midlife are associated with cognitive performance and plasma amyloid level assessed 15–20 years later. Faster biological aging was linked to worse cognition across multiple domains and lower Aβ42/Aβ40 ratios, whereas slower aging was associated with cognitive resilience. Longitudinal epigenetic aging trajectories may serve as early indicators of differential brain health risk even in midlife, when interventions to modify disease trajectories may be most effective.

## Data Availability

All data produced in the present study are available upon reasonable request through the CARDIA Coordinating Center.

https://sites.uab.edu/cardia/

## Acknowledgements

The Coronary Artery Risk Development in Young Adults Study (CARDIA) is conducted and supported by the National Heart, Lung, and Blood Institute (NHLBI) in collaboration with the University of Alabama at Birmingham (75N92023D00002 & 75N92023D00005), Northwestern University (75N92023D00004), University of Minnesota (75N92023D00006), and Kaiser Foundation Research Institute (75N92023D00003). This manuscript has been reviewed by CARDIA for scientific content.

Additional support was provided by the National Institute on Aging (NIA) through grants R01AG091431 and R35AG071916 (PI: Yaffe).

