## Supplements for "Epigenetic Clock Trajectories and Brain Health in Midlife"

**Supplementary Tables**

| **Outcome** | **Adj.R^2^** | **ΔR^2^** | **RMSE** | **F** | **df** | **p** | **N** |
| --- | --- | --- | --- | --- | --- | --- | --- |
| Aβ42/40 | 0.083 | 0.008 | 0.951 | 5.19 | (2, 1154) | 0.00571 | 1164 |
| NfL | 0.140 | <0.001 | 0.876 | 0.12 | (2, 1157) | 0.891 | 1167 |
| pTau217 | 0.134 | 0.001 | 0.893 | 0.74 | (2, 1156) | 0.479 | 1166 |
| DSST | 0.279 | 0.007 | 0.843 | 11.11 | (2, 2200) | 1.59E-05 | 2209 |
| MoCA | 0.333 | 0.006 | 0.793 | 9.39 | (2, 2192) | 8.73E-05 | 2201 |
| RAVLT | 0.235 | 0.007 | 0.869 | 10.42 | (2, 2202) | 3.13E-05 | 2211 |
| Stroop (reversed) | 0.154 | 0.003 | 0.906 | 3.57 | (2, 2190) | 0.0283 | 2199 |
| Total Verbal Fluency | 0.186 | 0.001 | 0.885 | 1.1 | (2, 2206) | 0.332 | 2215 |

**Supplementary Table 1.** **Model fit and incremental contribution of epigenetic aging trajectory group for cognitive and plasma AD Biomarker outcomes.** Each row is a separate model regressing the outcome on epigenetic aging trajectory group (±1 SD) and covariates: age, sex, race, education, physical activity, and *APOE* ε4 carrier status; biomarker models were additionally adjusted for eGFR. Adj. R² = adjusted R² of the full model (covariates + trajectory group); ΔR^2^ = increase in unadjusted R^2^ when trajectory group is added to the covariate-only model; RMSE = root mean square error (residual standard error) of the full model; F = partial F-statistic testing the trajectory group increment (full model vs. covariate-only reduced model), with degrees of freedom (numerator, denominator) shown in the df column; p = p-value for this partial F-test. N = number of participants. Cognitive outcomes are z-scored; biomarkers are log₁₀-transformed and z-scored.

**Supplementary Figures**

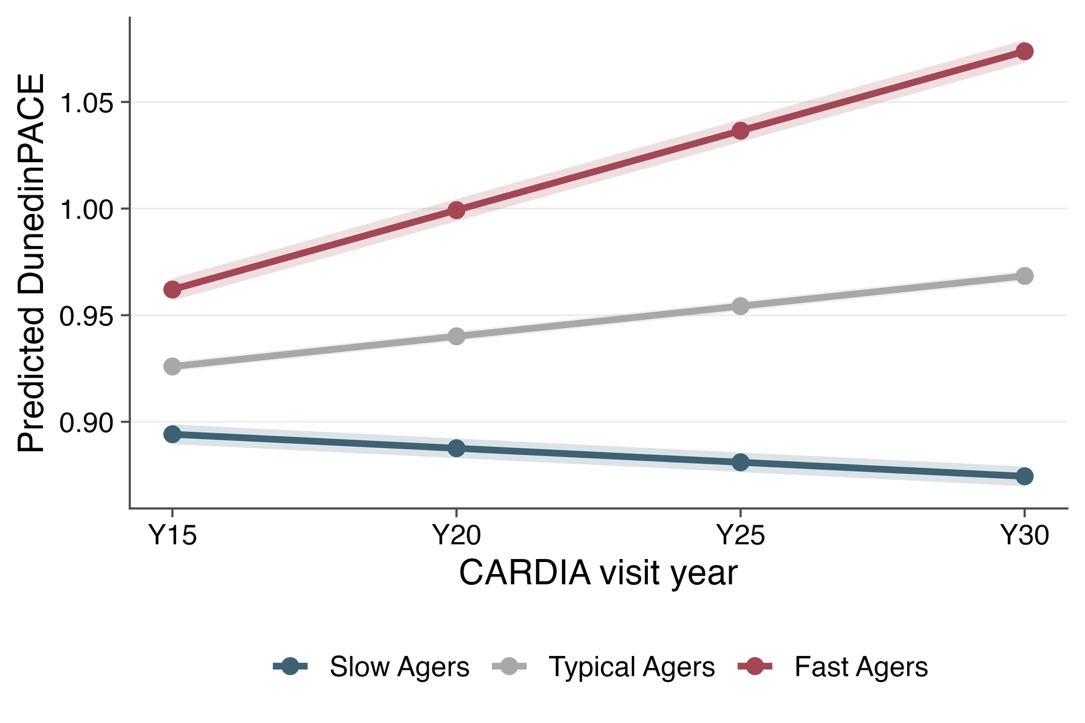

**Supplementary Figure 1. Predicted DunedinPACE trajectories by epigenetic aging group.** Model-predicted DunedinPACE (mean ± SD) trajectories across CARDIA Years 15–30 for each aging group, derived from a linear mixed-effects model with random intercepts and random slopes (time coded as years since baseline). Lines show group mean predicted values and shaded bands the standard error. Participants were classified by their individual random slopes as Slow Agers (blue), Typical Agers (gray), or Fast Agers (red). Fast Agers show the steepest increase in DunedinPACE over time, Typical Agers an intermediate increase at approximately the cohort-average rate, and Slow Agers a decline, illustrating the divergence in epigenetic aging rate used to define the trajectory groups.

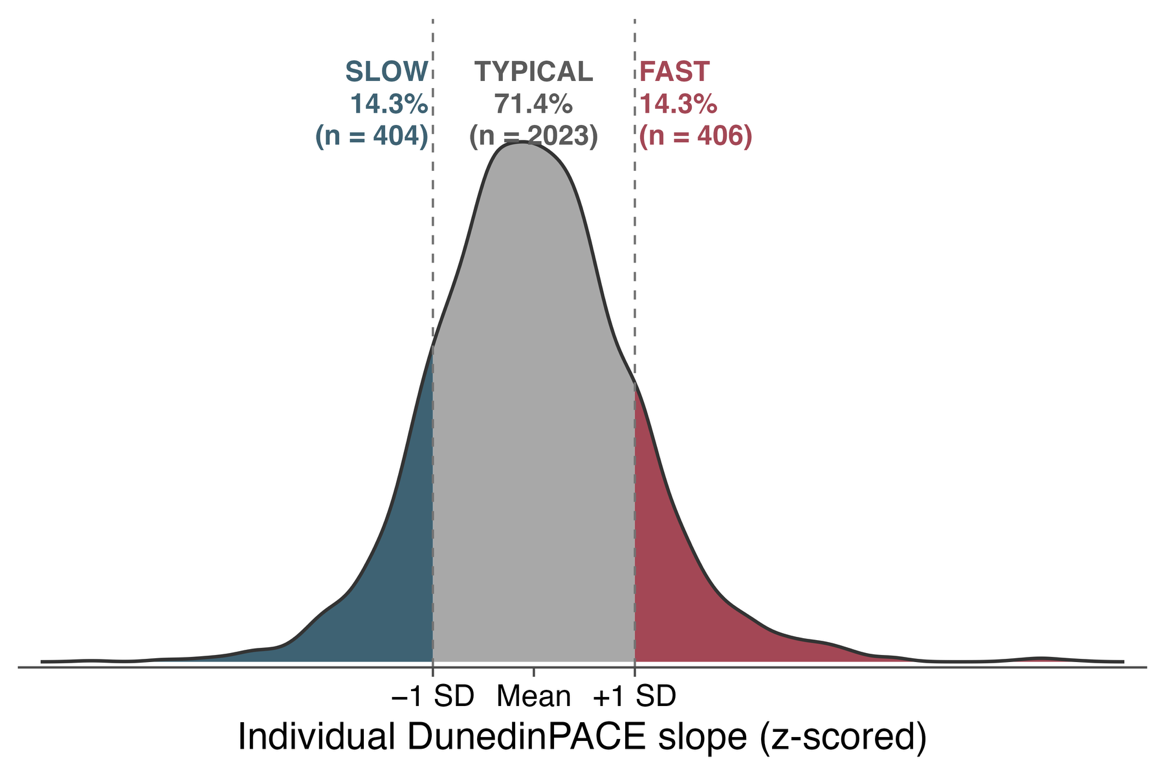

**Supplementary Figure 2. Distribution of individual epigenetic aging slopes and trajectory-group cut points.** Density of individual DunedinPACE slopes (z-scored) extracted from the linear mixed-effects model (N = 2,833). Dashed lines mark the ±1 SD cut points used for categorization. Participants were classified as Slow Agers (slope < −1 SD; n = 404, 14.3%), Typical Agers (within ±1 SD; n = 2,023, 71.4%), or Fast Agers (slope > +1 SD; n = 406, 14.3%).

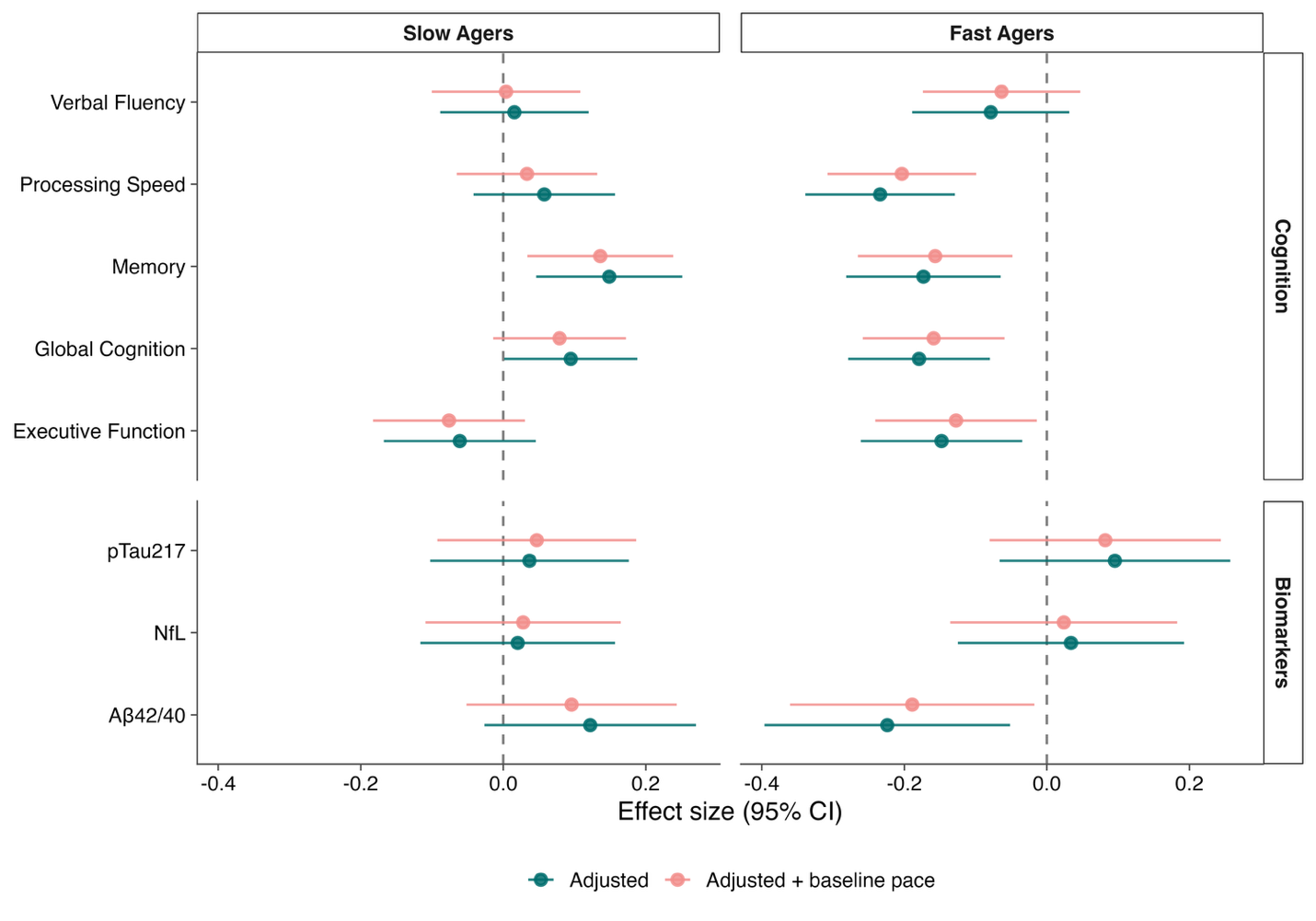

**Supplementary Figure 3:** **Sensitivity analysis: aging trajectory associations with additional adjustment for baseline epigenetic aging.** Point estimates (β) with 95% confidence intervals comparing Slow Agers and Fast Agers to Typical Agers (reference) for cognitive and plasma AD biomarker outcomes. Teal points show the primary adjusted model (age, sex, race, education, physical activity, and *APOE* ε4 carrier status; biomarker models additionally adjusted for eGFR); coral points show the same models with additional adjustment for baseline epigenetic aging, defined as the model-estimated DunedinPACE at Year 15 (sum of the fixed and participant-specific random intercepts from the trajectory model). Cognitive outcomes are z-scored; biomarkers are log₁₀-transformed and z-scored.
